# Imputation of race and ethnicity categories using continental genetic ancestry from real-world genomic testing data

**DOI:** 10.1101/2023.08.04.23293679

**Authors:** Brooke Rhead, Paige E. Haffener, Yannick Pouliot, Francisco M. De La Vega

## Abstract

The incompleteness of race and ethnicity information in real-world data (RWD) hampers its utility in promoting healthcare equity. This study introduces two methods—one heuristic and the other machine learning-based—to impute race and ethnicity from continental genetic ancestry using tumor profiling data. Analyzing de-identified data from over 100,000 cancer patients sequenced with the Tempus xT panel, we demonstrate that both methods outperform existing geolocation and surname-based methods, with the machine learning approach achieving high recall (range: 0.783-0.997) and precision (range: 0.913-0.981) across four mutually exclusive race and ethnicity categories. This work presents a novel pathway to enhance RWD utility in studying racial disparities in healthcare.

## 1. Introduction

Real-world data (RWD) offers insights into disease etiology, therapy outcomes, and racial disparities in healthcare.^1,2^ However, its utility in improving healthcare equity is limited by the significant sparsity of race and ethnicity data.^3,4^ This gap, attributable to factors such as lack of capture, data loss during transfer and de-identification, and shortcomings in electronic health record integrations,^5^ leads to reliance on limited, potentially biased datasets that may result in poorly generalizable results and biased disease outcome predictors.^4^

Several remediation strategies have been proposed, including improving data collection, conducting complete case analysis, modeling missingness in analyses, supplementing with additional data, and employing imputation methodologies.^6^ Existing imputation methods, many of which leverage census data based on geolocation and correlations between people’s surnames and their self-reported race and ethnicity,^7,8^ achieve moderate accuracy and require access to protected health information (PHI), limiting their applicability.^9^

Molecular tumor profiling, used for therapy decision support in cancer patients, generates a wealth of multimodal RWD that, once de-identified, can be harnessed for research.^10^ This type of RWD encompasses a wide array of clinical data as well as molecular and genetic data on a set of cancer-related and ancillary genes.^11^

Genetic ancestry inferred from sequencing data of molecular testing offers a potential solution to the challenge of race and ethnicity data missingness. The granularity of such inference is contingent on the availability of allele frequency data across reference population samples, with the most common level of genetic ancestry inference being at continental level categories, as described by the 1000 Genomes Project.^12^ Although genetic ancestry is not equivalent to race or ethnicity, a strong correlation between these two concepts has been observed among US populations.^13,14^ We propose to leverage this correlation and the genetic information available in molecular testing RWD using two methods — one heuristic and the other based on machine learning — to impute mutually-exclusive race and ethnicity categories from continental genetic ancestry. These methods were benchmarked and shown to outperform previously reported imputation methods, with the machine learning-based method providing the most accurate imputation.

## 2. Methods

The categorization of race and ethnicity in this study adheres to the standards developed by the US Office of Management and Business,^15^ which are also used in the US census. These standards are based on two self-reported questions: a) Race (American Indian or Alaska Native, Asian, Black or African American, Native Hawaiian or Other Pacific Islander, and White); and b) Ethnicity (Hispanic or Latino and Not Hispanic or Latino). However, these categories present analytical challenges due to the orthogonal race and ethnicity questions, and it is often more practical to consolidate answers to these two questions into non-overlapping classes,^16^ defined in this study as: Hispanic or Latino, non-Hispanic (NH) Asian, NH Black, and NH White, with the other races having insufficient numbers at the moment to develop reliable models in our source data. This consolidation allows for a more streamlined and comprehensive analysis of race and ethnicity in the context of RWD.

### 2.1. Data

Genomic and clinical data from patients of multiple cancer diagnoses was obtained from the Tempus database. The selected cohort consisted of 132,523 de-identified records of patients whose tissues were sequenced with the Tempus xT next-generation sequencing (NGS) panel (596-648 genes, v2-v4, tumor-normal matched when tissue available)^11,17^ from 2018 to 2022. A total of 31,491 had available race, ethnicity, and geolocation data and belonged to one of the four non-overlapping race and ethnicity categories that we imputed: 2,616 Hispanic or Latino, 1,258 NH Asian, 3,120 NH Black, and 24,497 NH White. An additional 62,674 records in our cohort had no available race or ethnicity data and were used as a comparison group. Analysis was performed using de-identified data under human subjects research exemption granted by Advarra, Inc. Institutional Review Board, protocol Pro00042950.

### 2.2. Determination of genetic ancestry

We estimated continental genetic ancestry proportions using a supervised global genetic ancestry estimation algorithm.^18^ This approach calculated the proportions for five continental ancestries — Africa (AFR), the Americas (AMR), East Asia (EAS), Europe (EUR), and South Asia (SAS)— using a bespoke set of 654 ancestry informative markers (AIMs) that intersect with the targeted regions of the Tempus xT NGS assay.^11^ We sourced reference allele frequency data for these AIMs from the 1,000 Genomes Project,^12^ the Human Genome Diversity Project,^19^ and the Simons Genome Diversity Project databases.^20^ To evaluate the accuracy of our methods, we compared our results with published ancestry proportions determined using the gold standard method, RFMix,^21^ on whole-genome sequencing data from the Pan-Cancer Analysis of Whole Genomes Project,^22^ showing an average normalized mean squared error of 0.12.

### 2.3. Benchmarking and performance metrics for race/ethnicity imputation

We relied on our cohort’s stated race and ethnicity data as specified in the Tempus database as our ground truth. To assess the performance of imputation methods, we employed a range of accuracy measures specific to each predicted race or ethnicity category. *Recall*, also called *sensitivity* or *true positive rate*,^*23*^ measures the proportion of individuals correctly assigned to a category among all individuals truly in that category. *Precision*, or *positive predictive value*,^*23*^ is the fraction of relevant instances among the retrieved instances; i.e., the proportion of correctly assigned individuals among all those assigned to a category. The *F1-score* is the harmonic mean of precision and recall, providing a balance between these two metrics. We also evaluated several measures of overall accuracy. *Cohen’s kappa*^*24*^ is a measure of agreement between predicted and true categories, accounting for the possibility of agreement occurring by chance. The *correct rate*, or *accuracy*,^*23*^ measures the proportion of all predictions that are correct. *Log loss* quantifies the difference between predicted probabilities of belonging to a class and the true value (0 or 1) of belonging to that class, with lower log loss indicating better model performance. The area under the receiver operating characteristic curve, or *AUC*, is a measure of model performance based on sensitivity and specificity across all classification thresholds and thus is not sensitive to any specific chosen threshold. *prAUC* is an analogous measure based on precision and recall.

In addition to these common measures, we also utilized metrics proposed by Elliot *et al*.^*7*^ The *weighted error* compares the true prevalence of race/ethnicity in the validation dataset to the predicted prevalence, providing an indication of the overall error rate. The *weighted correlation* measures the weighted average correlation (calculated using vectors of indicators) between true race and ethnicity and imputed category for each of the four categories, with weights equal to true prevalence. Together, these metrics offer a comprehensive evaluation of the performance of our imputation methods.

### 2.4. Heuristic imputation of race and ethnicity

We imputed mutually-exclusive race and ethnicity categories from continental genetic ancestry proportions using a set of heuristics (Table 1) derived from admixture proportions reported in the literature for Black and Hispanic/Latino groups in the United States.^14^ We defined four categories: NH Asian, NH Black, Hispanic/Latino, and NH White. Patients who did not fit heuristics’ categories were labeled “complex.”

**Table 1.** Race and ethnicity imputation heuristics from continental genetic ancestry. Continental ancestries codes: AFR, Africa; AMR, Americas; EAS, East Asia; EUR, Europe; SAS, South Asia.

| Imputed category | Continental genetic ancestry thresholds |
| --- | --- |
| Non-Hispanic Asian | >70% combined EAS and SAS |
| Non-Hispanic Black | >20% AFR, <10% AMR, and >70% combined AFR and EUR |
| Non-Hispanic White | >80% EUR and <10% AMR |
| Hispanic/Latino | >10% AMR and >70% combined AMR, EUR, and AFR |
| Complex | Remaining patients not meeting above thresholds |

### 2.5. Machine learning imputation of race and ethnicity

For all models, train+test and validation sets were assembled from a core dataset containing stated race, stated ethnicity, continental genetic ancestry proportions for AFR, AMR, EAS, EUR, and SAS, US census divisions (nine geographic groupings of states defined by the US Census Bureau), and demographic proportions for NH Asian, NH Black, NH White, and Hispanic or Latino of each patient’s ZIP code tabulation area (ZCTA), obtained from the 2021 5-year American Community Survey and mapped to three-digit ZIP codes using UDS Mapper.^25^ We tested three models using different types of features: 1) *ML-ancestry*: continental genetic ancestry proportions only, 2) *ML-ancestry+geolocation*: continental genetic ancestry proportions and US census divisions, 3) *ML-ancestry-demographic*s: continental genetic ancestry proportions and demographic proportions.

We implemented all machine learning models in R using the caret package (v 6.0.94)^26^ We initially considered models that implemented random forest (method=“rf”) and gradient boosting (method=“gbm”) algorithms. We ultimately chose a boosted logistic regression algorithm (method=“LogitBoost”) as it provided the ability to make “no-call” assignments with a probabilistic threshold (*p* ≥ 0.5) in classification. We split the train+test and validation sets 90/10 while maintaining US census division proportions in each set to ensure that the sets were aligned well for populations whose continental genetic ancestry proportions vary by U.S. geography, e.g., Hispanic or Latino.^14^ All models were trained using 10-fold cross validation. Grid expansion was performed to evaluate boosting iterations from 1 to 100 in intervals of 10. The optimal number of iterations and the final model were selected based on the lowest log loss value.

## 3. Results

### 3.1. Comparison of performance of race and ethnicity imputation methods

Table 2 summarizes the overall performance of the heuristic assignment method and each of the ML models. The ML model that solely considered continental genetic ancestry proportions achieved the best mean F1-score (0.937), Cohen’s kappa (0.919), correct rate (0.970), weighted correlation (0.914), and log loss (0.140), whereas the heuristic method performed the worst by most metrics: mean F1-score 0.923, Cohen’s kappa 0.894, correct rate 0.959, and weighted correlation 0.868. The ML model that added demographics in the form of the proportion of each race or ethnicity category residing in a patient’s three-digit ZCTA performed second-best for most metrics aside from Cohen’s kappa (third best) and AUC (best), while the ML model that included geolocation in the form of US Census district of a patient’s home address state performed third-best by most metrics except Cohen’s kappa (second best), log loss (tied for best), and prAUC (best).

**Table 2.** Overall performance of race and ethnicity imputation methods on validation set. Refer to section 2.5 for ML method descriptions. Best performing metric indicated with bold.

| Imputation Method | Mean<br>F1-Score | Cohen’s<br>Kappa | Correct<br>Rate | Weighted<br>Correlation | Weighted<br>Error | Log<br>Loss | AUC | prAUC |
| --- | --- | --- | --- | --- | --- | --- | --- | --- |
| Heuristic | 0.923 | 0.894 | 0.959 | 0.868 | **0.007** | - | - | - |
| ML (ancestry) | **0.937** | **0.919** | **0.970** | **0.914** | 0.008 | **0.140** | 0.973 | 0.924 |
| ML (ancestry + geolocation) | 0.933 | 0.915 | 0.968 | 0.912 | 0.011 | 0.143 | **0.975** | 0.713 |
| ML (ancestry + demographics) | 0.934 | 0.914 | 0.968 | 0.913 | 0.008 | **0.140** | **0.975** | **0.931** |

When evaluating performance by category, we found that recall, precision, and F1-score were all at or above 0.926 for the NH Asian, NH Black and NH White categories (Table 3). Performance of all imputation methods was worst for the Hispanic or Latino category, with recall ranging from 0.752-0.804, precision from 0.786-0.922, and F1-score from 0.795-0.843.

**Table 3.** Performance of race and ethnicity imputation methods on validation set per classification category. Refer to section 2.5 for ML method descriptions. Best performing metric for each category indicated with bold.

|  |  | Classification Category, N |  |  |  |
| --- | --- | --- | --- | --- | --- |
| Imputation Method | Metric | Hispanic or<br>Latino, 280 | NH Asian,<br>129 | NH Black,<br>304 | NH White,<br>2,433 |
| Heuristic | Recall | **0.804** | **0.992** | 0.974 | 0.973 |
|  | Precision | 0.786 | **0.947** | 0.931 | **0.983** |
|  | F1-Score | 0.795 | **0.969** | 0.952 | 0.978 |
| ML-ancestry | Recall | 0.783 | 0.969 | 0.997 | 0.987 |
|  | Precision | 0.913 | 0.939 | 0.938 | 0.981 |
|  | F1-Score | **0.843** | 0.953 | 0.966 | **0.984** |
| ML-ancestry + geolocation | Recall | 0.752 | 0.969 | **1.000** | **0.988** |
|  | Precision | **0.922** | 0.940 | 0.938 | 0.978 |
|  | F1-Score | 0.828 | 0.954 | **0.968** | 0.983 |
| ML-ancestry + demographics | Recall | 0.766 | 0.984 | 0.993 | 0.986 |
|  | Precision | 0.909 | 0.926 | **0.941** | 0.979 |
|  | F1-Score | 0.831 | 0.955 | 0.966 | 0.983 |

### 3.2. Performance of heuristic method

Perhaps unsurprisingly, the heuristic method for assigning race and ethnicity categories based on continental genetic ancestry proportions alone underperformed by nearly all measures as compared to all of the ML models. For the Hispanic or Latino category (the most difficult to predict using our input features), the heuristic method did have the highest recall (0.804), but this was achieved at the cost of low precision (0.786), also reflected in this method obtaining the lowest F1-score (0.795) for that category. The heuristic method did achieve the highest recall, precision, and F1-score in the NH Asian category. Overall, although the heuristic method did not perform as well as the ML method, its performance was not far behind, achieving an overall correct classification rate of 95.9%.

### 3.3. Performance of boosted logistic regression method

We found that the best performing boosted logistic regression model by most metrics was *ML-ancestry*. This model improved upon the heuristic method for all overall performance metrics, with an overall correct classification rate of 97.0%. It had lower recall (0.783) but higher precision (0.913) for the Hispanic or Latino category than the heuristic method, along with the highest F1-score for that category (0.843). The model had a recall of 0.969-0997 for the three non-Hispanic categories, indicating that it correctly identifies the vast majority of individuals in those categories and is usually correct in its predictions, with precision ranging from 0.938-0.981.

### 3.4. Performance of ML models including geolocation and demographics

Surprisingly, including geolocation or demographic composition from patients’ home address ZCTA areas to the continental genetic ancestry proportions (*ML-ancestry+geolocation*, and *ML-ancestry+demographics*) did not improve model performance according to most metrics, and in fact slightly worsened some overall performance metrics, yielding a correct classification rate of 96.8% for both models. The *ML-ancestry+geolocation* model did have a high AUC (0.975), but low prAUC (0.713), while the *ML-ancestry+demographics* model had the same high AUC (0.975) and the highest prAUC (0.931). Individual category performance metrics followed a similar pattern to the *ML-ancestry* model. Notably, the *ML-ancestry+geolocation* model had the best precision for the Hispanic or Latino category (0.922), which may be desirable for use cases where correct predictions are valued over high recall.

### 3.5. Reclassification of stated race/ethnicity categories by imputation

To gain deeper insights into the classification ability of the *ML-ancestry* model, we applied the model to the entire labeled dataset (train+test and validation). Table 4 presents a confusion matrix comparing the imputed categories with the race and ethnicity data from the Tempus database, including the rate of no-calls and the number and fraction of misclassified subjects. The confusion matrix for the validation dataset mirrors this table in terms of percentages (data not shown). Additionally, Figure 1 provides a visual representation of the allocation of subjects from their stated race and ethnicity to their imputed categories through a flow diagram.

**Table 4.** Confusion matrix comparing imputed race and ethnicity category to stated category for *ML-ancestry* model on the full labeled dataset. Percentage and numbers of subjects (in parentheses) are indicated in each cell.

| Imputed category | Stated category |  |  |  |
| --- | --- | --- | --- | --- |
|  | Hispanic or Latino | NH Asian | NH Black | NH White |
| Hispanic or Latino | 77.9% (2,037) | 0% (0) | 0.2% (6) | 0.7% (169) |
| NH Asian | 1.1% (28) | 95.8% (1,205) | 0.1% (4) | 0.2% (46) |
| NH Black | 2.4% (63) | 0.2% (3) | 98.8% (3,082) | 0.2 % (57) |
| NH White | 13.7% (359) | 2.4% (30) | 0.5% (15) | 98.4% (24,100) |
| No Call | 4.9% (129) | 1.6% (20) | 0.4% (13) | 0.5% (125) |
| Misclassified | 17.2% (450) | 2.6% (33) | 0.8% (25) | 1.1% (272) |

**Fig. 1.**
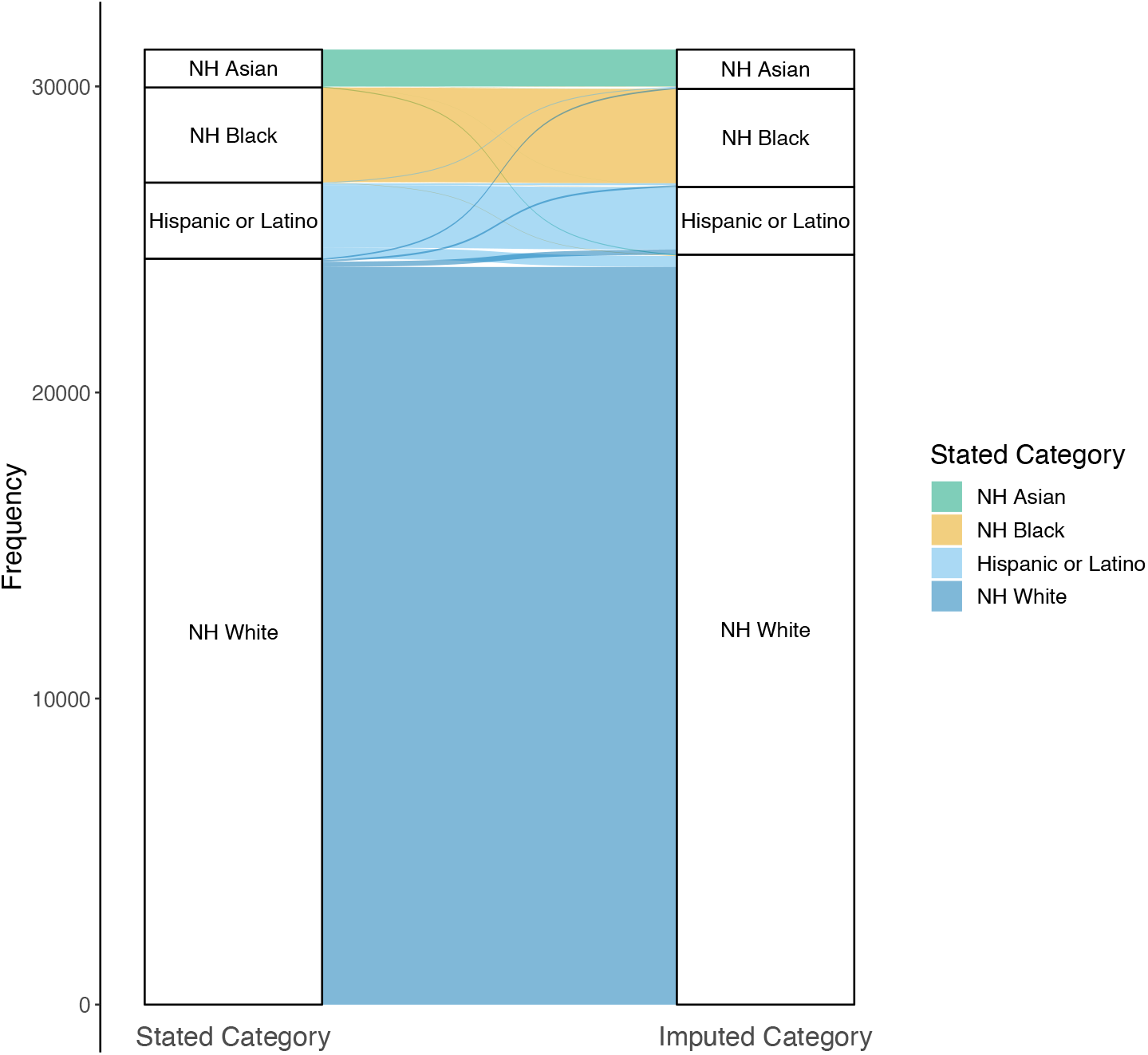
Flow diagram showing the relationship between stated (left) and imputed (right) race/ethnicity categories with the *ML-ancestry* model in the full labeled (train+test and validation) dataset of 31,491 de-identified patients.

The confusion matrix in Table 4 indicates that the Hispanic or Latino category experiences the highest rate of no-calls and misclassifications, whereas the NH Black category has the lowest. The flow diagram in Figure 1 emphasizes that most subjects are assigned to their stated category, with the majority of misclassifications occurring between Hispanic or Latino and NH White categories. This underscores the notion that ethnicity, being more of a cultural and/or country of origin construct, doesn’t align as closely with continental genetic similarity as the race categories do.^13^

### 3.6. Distribution of race and ethnicity categories imputed on unlabeled data

We also imputed race and ethnicity categories on a subset of 62,674 individuals with no available race or ethnicity data (“unlabeled”) using the *ML-ancestry* model. Compared to imputed categories for those with stated race and ethnicity (“labeled”), there are slightly more NH Asians, Hispanic/Latinos and No Calls, and fewer NH Whites (Figure 2).

**Fig. 2.**
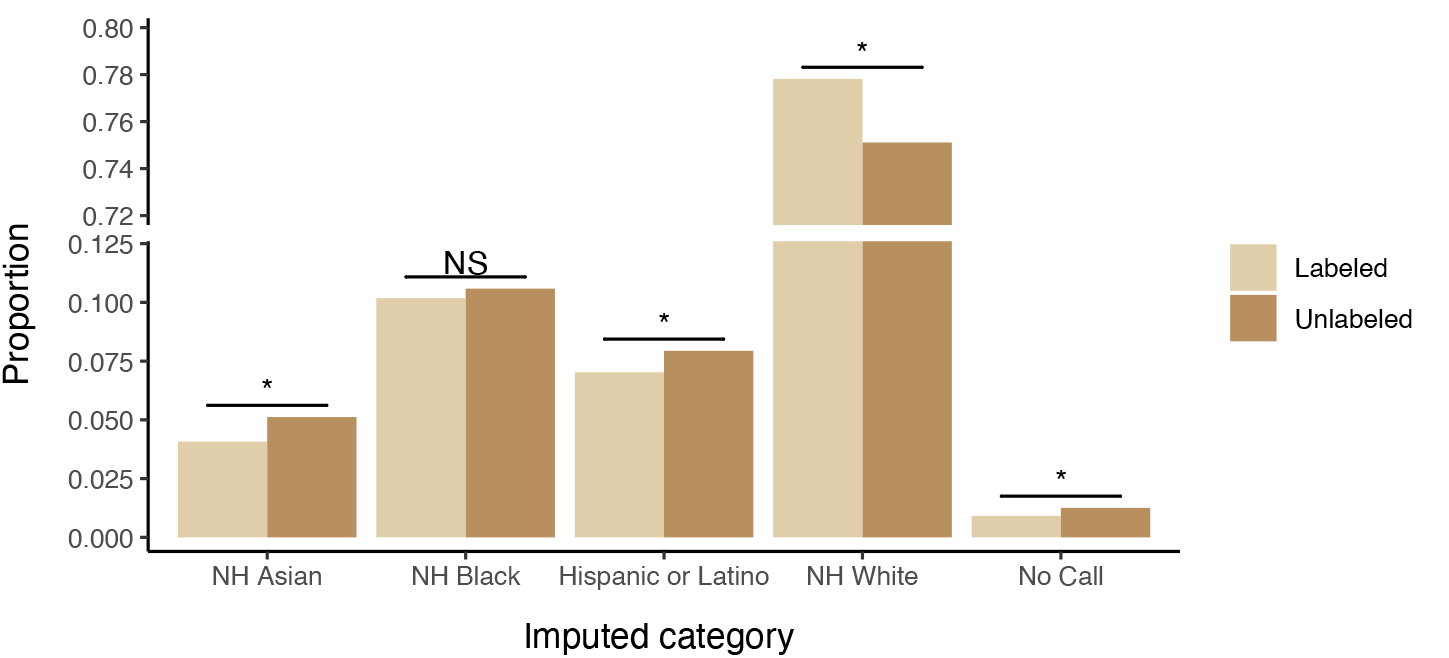
Proportions of race and ethnicity categories imputed using the *ML-ancestry* model in the labeled (N=31,491) and unlabeled (N=62,674) datasets. Stars indicate a significant (Bonferroni-corrected p<0.05) difference in proportions in a post hoc analysis of standardized residuals from a χ^2^ test of independence. NS, not significant. Note the break in the y-axis.

### 3.7. Analysis of potential biases

The dataset used to develop our ML models is heavily imbalanced, with the largest group of patients (∼78%) having a stated category of NH White, and the smallest group (∼4%) having a stated category of NH Asian, potentially leading to overfitting to the majority category and biasing model performance. To address these potential problems, we evaluated additional models in parallel to those discussed here, wherein each model was trained in the same way except that each train+test set was downsampled to require an equal number of patients in each category, matching that of the category with the smallest number of patients. However, the downsampled models showed worse overall performance by all of our metrics and, within each category, had lower F1-scores (data not shown). Additionally, the performance metrics computed on the train+test sets during cross-validation were only slightly better than those computed on the validation set, alleviating concerns of overfitting. Importantly, the performance metrics we considered included those broken down by classification category to enable evaluation of whether any particular category was underperforming relative to the others. We also considered metrics that are suited to imbalanced data, such as Cohen’s kappa.

## 4. Discussion

While a direct comparison of the performance of our methods with other imputation methods was not possible in our dataset due to the unavailability of PHI such as surnames or addresses in a de-identified setting, we compared our performance with their reported performance in the literature suggesting that our method substantially outperforms these prior methods;^7–9^ e.g. the weighted correlation was at least 9% better than the BISG method,^8^ and the weighted error of all of our models are an order of magnitude lower than those studied by Xue *et al*.^*9*^

The category with the lowest recall was Hispanic or Latino, ranging from 75-80%, with the highest level of no-calls also observed for this category (5% vs 1-3%). However, the *ML-ancestry-geolocation* model provided the best precision (92%) at a good recall rate (75%), while the Heuristic method provided the best recall (80%) but at a significantly lower level of precision (79%). Although the intended use of the imputation may dictate the best trade-off, we believe that precision is the most important feature as minimization of misclassified subjects is generally more desirable. The drop in performance in the Hispanic or Latino category may be due to the fact that self-affiliation with this category corresponds more with culture and language than with genetic similarity, with levels of admixture within this group varying widely depending on country of origin and among the coasts of the US.^14^

Our work has potential limitations. Differences between patients with complete vs. incomplete stated race and ethnicity could affect training and therefore imputation performance. The unequal distribution of imputed categories in labeled and unlabeled data suggests that there are indeed some slight differences in the composition of patients who lack race and ethnicity data, with imputed NH Asian and Hispanic or Latino categories most likely to be missing this information, but therefore also most likely to benefit from imputation. Given the limited numbers of American Indian or Alaska Native and Native Hawaiian or Other Pacific Islander individuals in our dataset as well as the lack of public allele frequency information from these groups, we are unable to develop models to impute those categories, meaning those individuals will be misclassified, typically as Hispanic or Latino and NH Asian, respectively. As our database grows and additional AIM allele frequencies become available, our model could be retrained for these additional categories. While the performance of our models on populations outside the US is unknown, or indeed performance with differently ascertained population samples, our results suggest that retraining with additional data pertaining to those populations could yield similar performance in other settings.

When developing our race imputation methods, we adhered to established recommendations for ethical imputation.^27^ We audited input data for bias, scrutinized methodological choices for potential bias introduction, and rigorously assessed the accuracy of the imputed data. Our adherence to these guidelines underscores our commitment to the responsible use of race imputation in promoting equity in healthcare.

## 5. Conclusions

Addressing racial disparities is pivotal to advancing equity in precision medicine. However, the frequent unavailability of data disaggregated by race and ethnicity in RWD can lead to biased outcome predictors, inadequate representation in clinical trials, and poorly targeted policies, potentially exacerbating disparities. While complete self-reported data is the ultimate goal and could offer optimal race and ethnicity information, the associated costs, time, and potential ethical risks render it an impractical solution in some settings. Our study underscores the efficacy of using continental genetic ancestry data to impute race and ethnicity categories in a de-identified setting, effectively mitigating the challenge of data sparsity for these categories in RWD in the US. This approach could allow more accurate identification of racial disparities in certain healthcare settings where genetic data are available, contributing to the development of more targeted and equitable healthcare interventions.

## Data Availability

The datasets supporting the conclusions of this article are included within the article. Due to patient privacy, data sharing consent, and HIPAA regulations, our raw data cannot be submitted to publicly available databases.

## Acknowledgments

We appreciate the discussions and feedback received on this work from Funmi Olopade (University of Chicago), John Carpten (City of Hope), Jose Trevino (Virginia Commonwealth University), Carlos D. Bustamante (Stanford University), Calvin Chao, and Ezra Cohen (Tempus Labs). We also acknowledge Frank Nothaft, Rafael Esleyer, Nick Riggan, and Arvind Prasad from the Tempus Lens team for their assistance in procuring de-identified data and correcting data problems needed for this work. We thank Vanessa Nepomuceno from the Tempus Publications team for copyediting the manuscript.

